# Exploring approaches to weighting estimates of facility readiness to provide health services used for estimating input-adjusted effective coverage: a case study using data from Tanzania

**DOI:** 10.1101/2023.03.30.23287947

**Authors:** Ashley Sheffel, Emily Carter, Debora Niyeha, Khadija I. Yahya-Malima, Deogratius Malamsha, Shagihilu Shagihilu, Melinda Munos

## Abstract

The ideal approach for calculating effective coverage of health services using ecological linking requires accounting for variability in facility readiness to provide health services and patient volume by incorporating adjustments for facility type into estimates of facility readiness and weighting facility readiness estimates by service-specific caseload. The aim of this study is to compare the ideal caseload-weighted facility readiness approach to two alternative approaches 1) facility-weighted readiness and 2) observation-weighted readiness to assess the suitability of each as a proxy for caseload-weighted facility readiness. We utilized the 2014-2015 Tanzania Service Provision Assessment along with routine health information system data to calculate facility readiness estimates using the three approaches. We then conducted equivalence testing, using the caseload-weighted estimates as the ideal approach and comparing with the facility-weighted estimates and observation-weighted estimates to test for equivalence. Comparing the facility-weighted readiness estimates to the caseload-weighted readiness estimates, we found 58% of estimates met the requirements for equivalence. In addition, the facility-weighted readiness estimates consistently underestimated, by a small percentage, facility readiness as compared to the caseload-weighted readiness estimates. Comparing the observation-weighted readiness estimates to the caseload-weighted readiness estimates, we found 64% of estimates met the requirements for equivalence. We found that, in this setting, both facility-weighted readiness and observation-weighted readiness may be reasonable proxies for caseload-weighted readiness. However, in a setting with more variability in facility readiness or larger differences in facility readiness between low caseload and high caseload facilities, the observation-weighted approach would be a better option than the facility-weighted approach. While the methods compared showed equivalence, our results suggest that selecting the best method for weighting readiness estimates will require assessing data availability alongside knowledge of the country context.

## INTRODUCTION

In low- and middle-income countries (LMICs), effective coverage estimates of health service provision are often generated by linking data from household surveys and health facility assessments (HFAs) (1). The ideal approach is exact-match linking whereby information for each individual seeking care in the household survey is linked to information about the quality of care of the specific health facility visited. However, this is generally not feasible for large-scale, national surveys in LMICs (2). An alternative is ecological linking, whereby each individual seeking health care services in the household survey is linked to an average quality of care score of health facilities within the same administrative area as the household (3-5).

Health facilities within an administrative area are often of varying types (hospitals, health centers, health posts) and as such they are often not equally ready to deliver care and they do not care for equal volumes of patients. It is important to account for this variability when generating linked estimates as it reduces bias and results in a closer approximation to an exact-match link. This can be accomplished by incorporating adjustments for facility type into estimates of facility readiness and by weighting facility readiness estimates by service-specific caseload (3-5).

Data on facility type is readily available in HFAs and incorporating adjustments for facility type into estimates of facility readiness is a recommended best practice for ecological linking. However, caseload data is not collected in standard HFA tools. It can also be difficult to obtain caseload data and link it to HFA data. The aim of this study is to use data from Tanzania to compare the ideal caseload-weighted facility readiness approach to two alternative approaches: 1) facility-weighted readiness and 2) HFA observation-weighted readiness, to assess the suitability of each approach as a proxy for caseload-weighted facility readiness.

## METHODS

### Data sources

#### Tanzania Service Provision Assessment 2014-2015 (TSPA)

The 2014-2015 TSPA was a health facility assessment that included a standard set of survey instruments: a facility inventory questionnaire, health worker interviews, observation of ANC consultations, and exit interviews with ANC clients. The survey was sampled to be nationally representative with health facilities selected using stratified systematic probability sampling with stratification by region and facility type (equal probability within strata) with oversampling of hospitals (see ***Box 1*** for details on facility types). The TSPA final report contains comprehensive information on the survey methodology and questionnaires (6). The TSPA dataset is publicly available from the DHS Program data repository but has been de-identified to exclude health facility names.

##### Box 1

Facility types in Tanzania (7)

Facility types in Tanzania include the following:

- **Dispensary** – Dispensaries are generally the first point of contact with the health care system and are staffed by a clinical assistant often with the support of a nurse. Services provided at dispensaries include maternal and child health care, assistance with uncomplicated deliveries, and basic outpatient curative care.
- **Health center** – Health centers supervise the dispensaries and are staffed by a clinical officer often with the assistance of a clinical assistant, maternal and child health aide, health aide, and a health assistant. Services provided at health centers include preventative care as well as curative care for common diseases and minor surgery.
- **Clinic** – Clinics are private primary-level health facilities which provide mostly outpatient curative services. They employ nurses/midwives, clinical officers, doctors, and pharmaceutical technicians.
- **Hospital** – This includes national referral hospitals, regional hospitals, district hospitals, and private hospitals. Secondary care is provided by district hospitals, which are staffed by medical doctors and assistant medical officers supported by clinical officers and nurses. District hospitals offer both inpatient and outpatient services not available at lower level facilities including laboratory, imaging, and surgical services. Tertiary care is provided at regional hospitals and national referral hospitals, which are staffed by surgeons and medical doctors, as well as general and specialized nurses and midwives. They offer similar services as district hospitals; however, they are larger, employ specialists in various fields, and offer additional advanced services.

#### TSPA Facility names dataset

The TSPA Facility names dataset contains the health facility names and unique IDs that can be linked to the 2014-2015 TSPA dataset. This data was made available by the Tanzania National Bureau of Statistics as part of the National Evaluation Platform (NEP) project implementation.

#### Tanzania Health Facility Registry

The Health Facility Registry (HFR) is a web-based system used to provide public access to a database of information about all health facilities in mainland Tanzania. The HFR registry contains the name, region, and facility ID for each health facility and can be accessed at https://hfrportal.moh.go.tz/.

#### Tanzania DHIS-2 service utilization data

Data on the number of ANC4+ visits (defined as a count of all ANC visits made that were the 4th or more visit for a woman) was extracted from the Tanzania DHIS-2 portal per month and per facility for 2015.

### Caseload estimates

The data on ANC4+ visits was exported from DHIS-2 for each region to a .csv file. All non-facility-level entries were dropped (i.e., district and region values), and duplicate entries were dropped. We then assessed the extent of missingness of ANC4+ data across the 12 months and created a binary variable with 0 indicating <=5 months of missing data and 1 indicating >5 months of missing data. Missing data was imputed with the facility-specific mean value of ANC4+ visits. ANC4+ caseload was calculated as the total number of ANC4+ visits across the 12-month period (January-December 2015).

### ANC readiness score

We defined facility readiness as the human resources, equipment and supplies, diagnostics, medicines, and basic amenities essential to deliver a high-quality ANC service. A total of 19 items from the TSPA were selected to include in the facility readiness index. Details on item selection have been previously published (8). For each item, a facility received one point if the requirements were met and a zero if not. A simple additive approach was utilized to calculate the index by taking the sum of the items available, dividing by the total number of items in the index, and multiplying by 100 to create a score between zero and 100.

### Linking TSPA and DHIS-2 data

We used an the HFR dataset to link the TSPA and DHIS-2 datasets. We merged the 2015 TSPA and DHIS-2 datasets using the HFR registry to identify facilities by both their name (TSPA) and facility identification code (DHIS-2)

### Calculate weighted facility readiness estimates

We aggregated estimates of facility quality over select categories of facility type, managing authority, and geographic unit aligning with different levels of aggregation that could be employed in ecological linking. Specifically, we generated aggregate quality scores by facility type, managing authority, urbanicity, and region. We limited the analysis to facilities that offered ANC services, had complete data for all facility readiness variables, and had at least 7 months of routine data available to calculate caseload. We weighted our aggregate facility readiness estimates using three approaches.

#### Approach 1: facility-weighted

The TSPA survey dataset includes survey weights (health facility weight, provider weight, client weight) that are calculated by the DHS program. The health facility weight is calculated based on the health facility selection probability, adjusted for non-response at the sampling stratum level. The facility-weighted approach used only the TSPA health facility weight to generate the weighted facility readiness estimate, scaling each facility’s contribution to align with the national distribution of facilities.

#### Approach 2: observation-weighted

For the observation-weighted approach, we approximated differentials in client volume by multiplying each facility’s weight by the number of client observations and the inverse probability of client selection. In addition, we limited the client observation dataset to women for whom facility level data was included in the analysis (i.e., in mainland Tanzania, offered ANC services, complete data for all facility readiness variables, and at least 7 months of routine data). The client observation dataset includes a client weight that is calculated by taking the health facility weight multiplied by the inverse selection probability of clients within each of the sampling strata, adjusted for client non-response. By scaling the facility weight by the inverse probability of client selection, the contribution of each facility’s readiness to the category average is adjusted to reflect the client load.

#### Approach 3: caseload-weighted

For the caseload-weighted approach, we multiplied the health facility weight by the ANC4+ caseload estimate to generate a caseload weight. We applied these caseload weights when calculating mean facility readiness by category.

### Analysis

All estimates were generated using the R “survey” package (9). In addition to specifying the weights for each approach, we also accounted for the HFA sampling design, including cluster (health facility) and stratification (facility type and region). We then conducted equivalence testing, using the caseload-weighted estimates as the ideal approach and comparing with the facility-weighted estimates and observation-weighted estimates to test for equivalence with an equivalence interval of (−5% to 5%). All statistical analyses were carried out using R version 4.1.3 (10).

## RESULTS

### Survey characteristics

The 2014-2015 TSPA data set contained data from 1078 health facilities in mainland Tanzania, of which 949 provided ANC services. All 949 facilities offering ANC services had complete data for all facility readiness variables, while 860 facilities had at least 7 months of routine data available to calculate caseload. On average, each facility had an annual caseload of 45 ANC4+ clients. The annual health facility ANC4+ caseload ranged from 2 to 891. There was a total of 3466 ANC client consultations from 689 facilities observed in the 2014-2015 TSPA. On average, each facility had 5 ANC client observations. The number of ANC client observations at a health facility ranged from 1 to 15.

### Variability in caseload

Caseload was variable both across and within facility type. Clinics and dispensaries tended to have lower caseload while health centers and hospitals tended to have comparatively higher caseload. However, within health centers and hospitals, there were facilities with both very low caseload and very high caseload (***Figure 1***).

**Figure 1:**
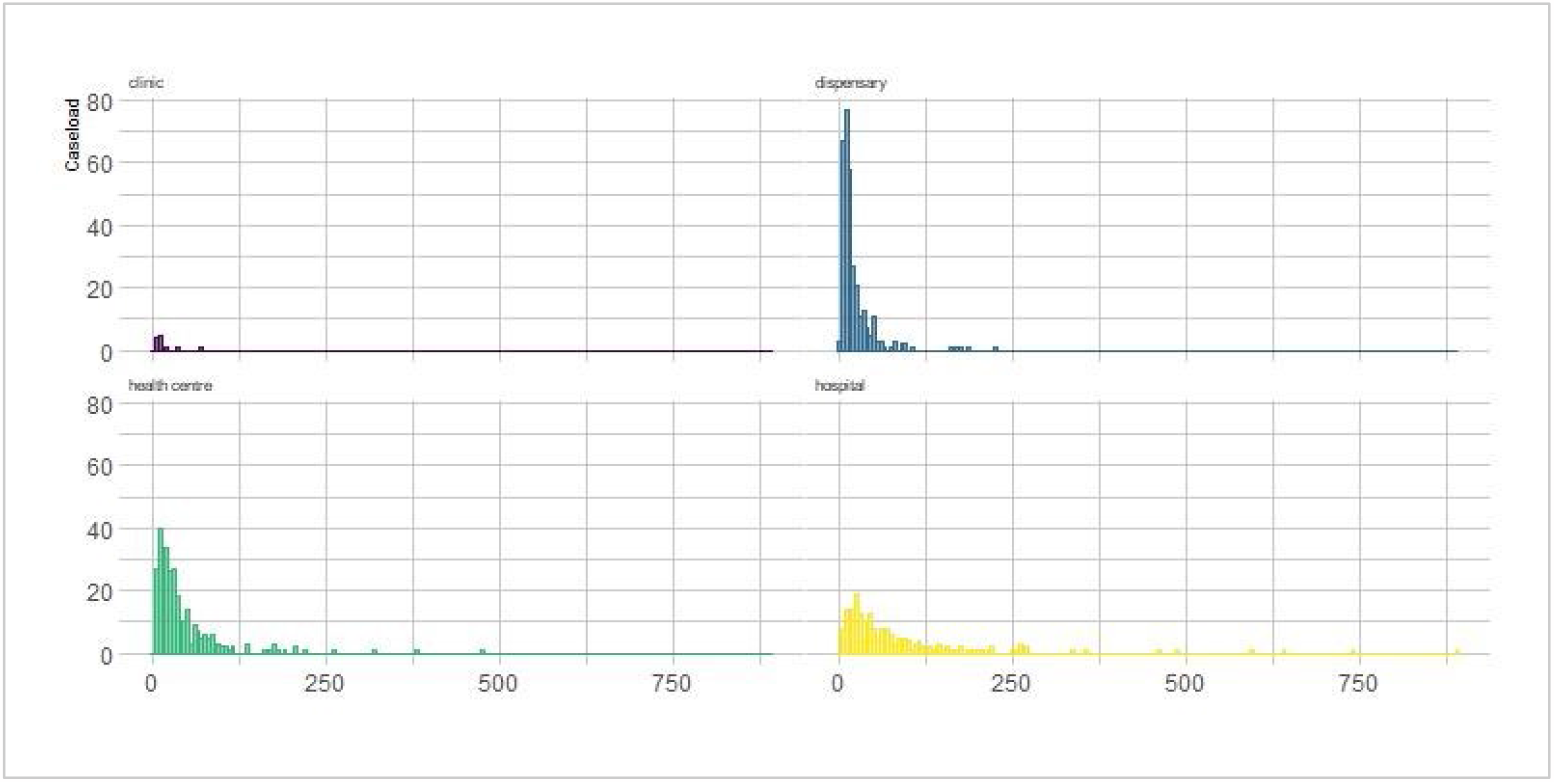
Histograms of caseload by facility type.

**Figure 2:**
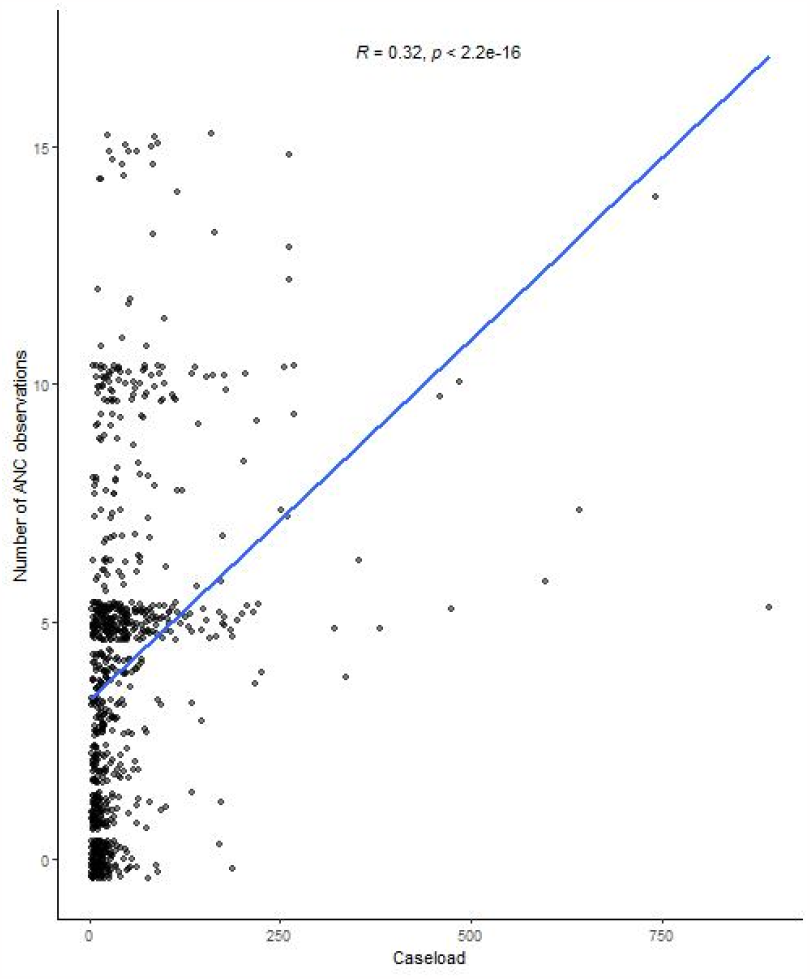
Correlation between caseload and number of ANC observations.

### Correlation between caseload and number of ANC observations

There was a weakly positive correlation (R = 0.32) between caseload and the number of ANC observations at a health facility.

### Facility readiness estimates and equivalence

The detailed results comparing the ideal caseload-weighted facility readiness approach to facility-weighted readiness and observation-weighted readiness are in ***Supplementary Table 1*** and a summary of the findings is in ***Table 1***.

**Table 1:** Summary of equivalence results.

| Equivalence finding | Caseload vs. Facility | Caseload vs. Observation |
| --- | --- | --- |
| Statistically significantly different, equivalent | 16 | 19 |
| Not statistically significantly different, equivalent | 5 | 4 |
| Statistically significantly different, not equivalent | 15 | 13 |

Comparing the facility-weighted readiness estimates to the caseload-weighted readiness estimates, we found 14% of the estimates were not statistically significantly different and were equivalent, which is what we would expect to see if the method was an exact approximation of the ideal caseload approach. An additional 44% of the estimates were statistically significantly different but were equivalent, which can be considered reasonable approximations of the ideal caseload approach. Combined, 58% of estimates met the requirements for equivalence, while 42% of estimates were statistically significantly different and not equivalent. The non-equivalent facility-weighted estimates were all regional estimates plus one managing authority estimate (parastatal). The facility-weighted readiness estimates consistently underestimated, by a small percentage, facility readiness as compared to the caseload-weighted readiness estimates with 33 out of 36 (92%) facility-weighted readiness estimates being lower than the caseload-weighted readiness estimates.

Comparing the observation-weighted readiness estimates to the caseload-weighted readiness estimates, 11% of the estimates were not statistically significantly different and were equivalent. An additional 53% of the estimates were statistically significantly different but were equivalent, for a total of 64% of estimates meeting the requirements for equivalence. Thirty six percent of the estimates were statistically significantly different and not equivalent. The non-equivalent observation-weighted estimates were all regional estimates. There was no consistent trend in overestimation or underestimation when comparing the observation-weighted readiness estimates to the caseload-weighted readiness estimates.

## DISCUSSION

We found that, in this setting, both facility-weighted readiness and observation-weighted readiness may be reasonable proxies for caseload-weighted readiness, as the national readiness estimates and readiness estimates disaggregated by facility level, urbanicity, and managing authority all met the criteria for equivalency (except for the parastatal estimate for facility-weighted readiness).

However, we found that the facility-weighted estimates consistently underestimated facility readiness, albeit by a small percentage, which occurred because facilities with higher readiness scores (which tended to have higher caseloads) were consistently upweighted in the caseload-weighted approach. In a setting with more variability in facility readiness or larger differences in facility readiness between low caseload and high caseload facilities, this underestimation could be more pronounced, and the observation-weighted approach would be a better option than the facility-weighted approach.

This study had a number of limitations. Data quality challenges with the routine data utilized for calculating caseload created an imperfect measure for our “ideal” approach. We addressed some of the data quality issues by imputing missing data and excluding facilities with more than 40% missing data. Data quality is often a challenge for routinely collected data in LMICs, so this analysis likely reflects what is possible to achieve in other similar contexts (11, 12). This study was performed using data from a single country, which may limit generalizability. However, this approach could easily be replicated in other contexts with caseload data that could be linked to health facility data.

This study has provided an important contribution to the growing evidence around best practices for generating effective coverage estimates. While the methods compared showed equivalence, our results suggest that selecting the best method for weighting readiness estimates will require assessing data availability alongside knowledge of the country context.

## Data Availability

All data produced in the present study are available upon reasonable request to the authors

## ACKNOWLEDGEMENTS

The authors wish to acknowledge the Bill & Melinda Gates Foundation for their support of this project.

## AUTHOR CONTRIBUTIONS

AS, EC, and MKM contributed to conceptualizing the paper and analysis. DM, DN, KYM, and SS performed data extraction and management. AS drafted the manuscript, with critical review and revision from all authors.

## DISCLOSURE STATEMENT

No potential conflict of interest was reported by the authors.

## ETHICS AND CONSENT

This is a secondary analysis and as such did not involve human subjects research.

## FUNDING INFORMATION

This work was supported by the Improving Measurement and Program Design grant (OPP1172551) from the Bill & Melinda Gates Foundation.

## PAPER CONTEXT

Effective coverage estimates, increasingly used to track progress toward universal health coverage, are often generated by linking data from household surveys and health facility assessments. However, little research has been done on approaches to weighting estimates of facility readiness used for estimating input-adjusted effective coverage. This paper provides a comparison of three methods for calculating weighted facility readiness estimates. Insights from this study serve to advance best practices in methodologies for generating effective coverage estimates.

## SUPPLEMENTARY MATERIAL

**Supplementary Table 1:**
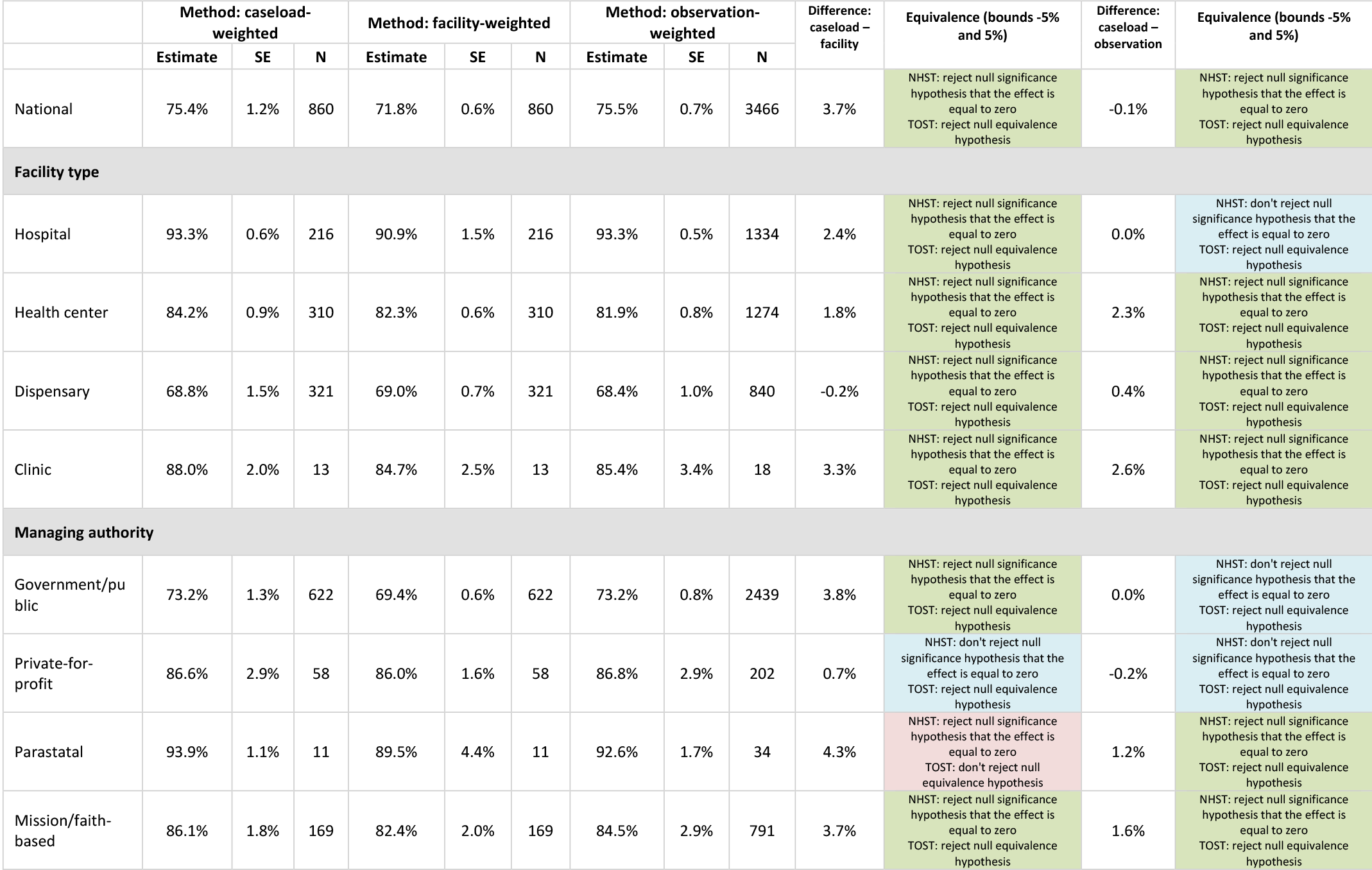
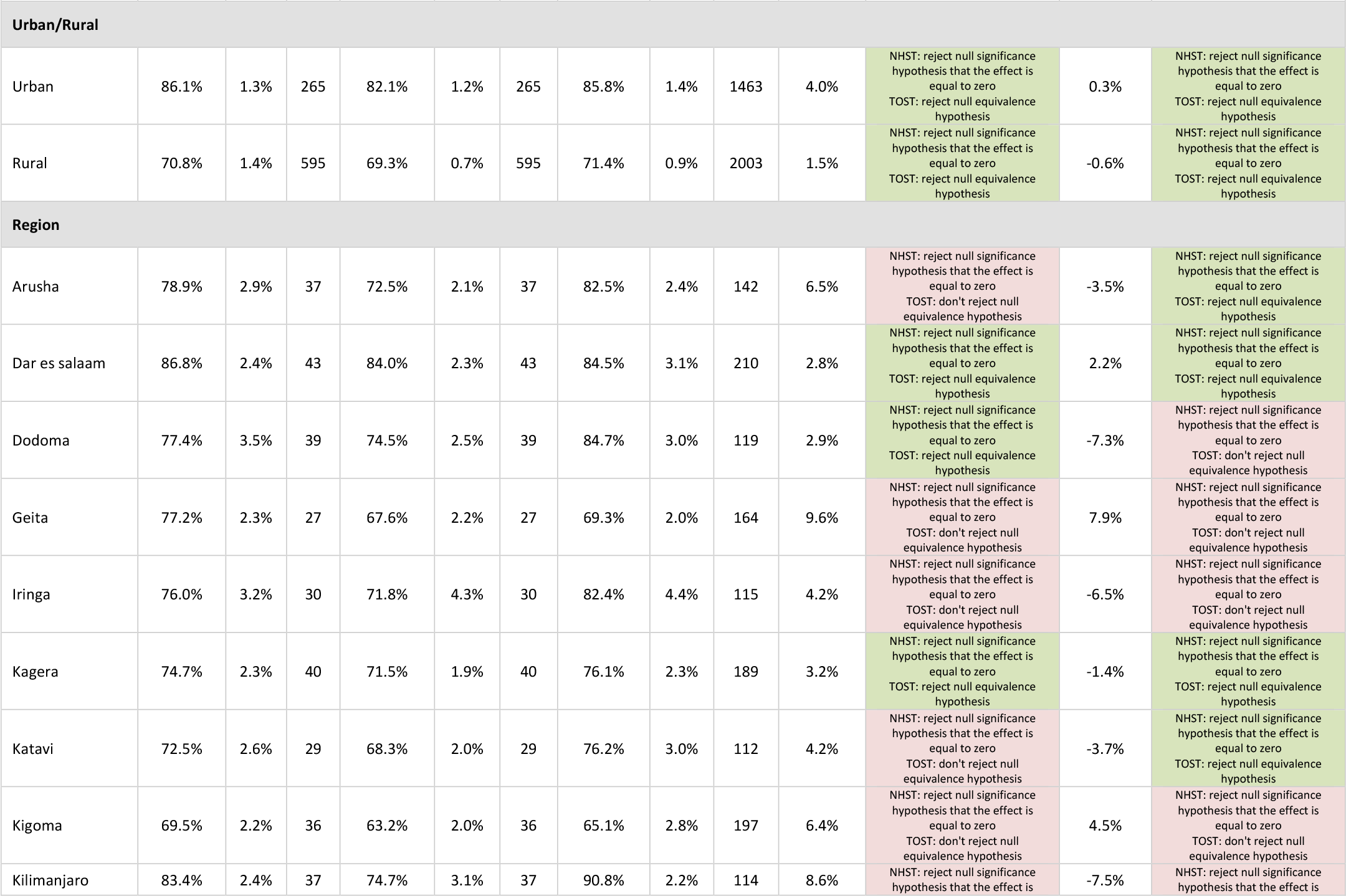
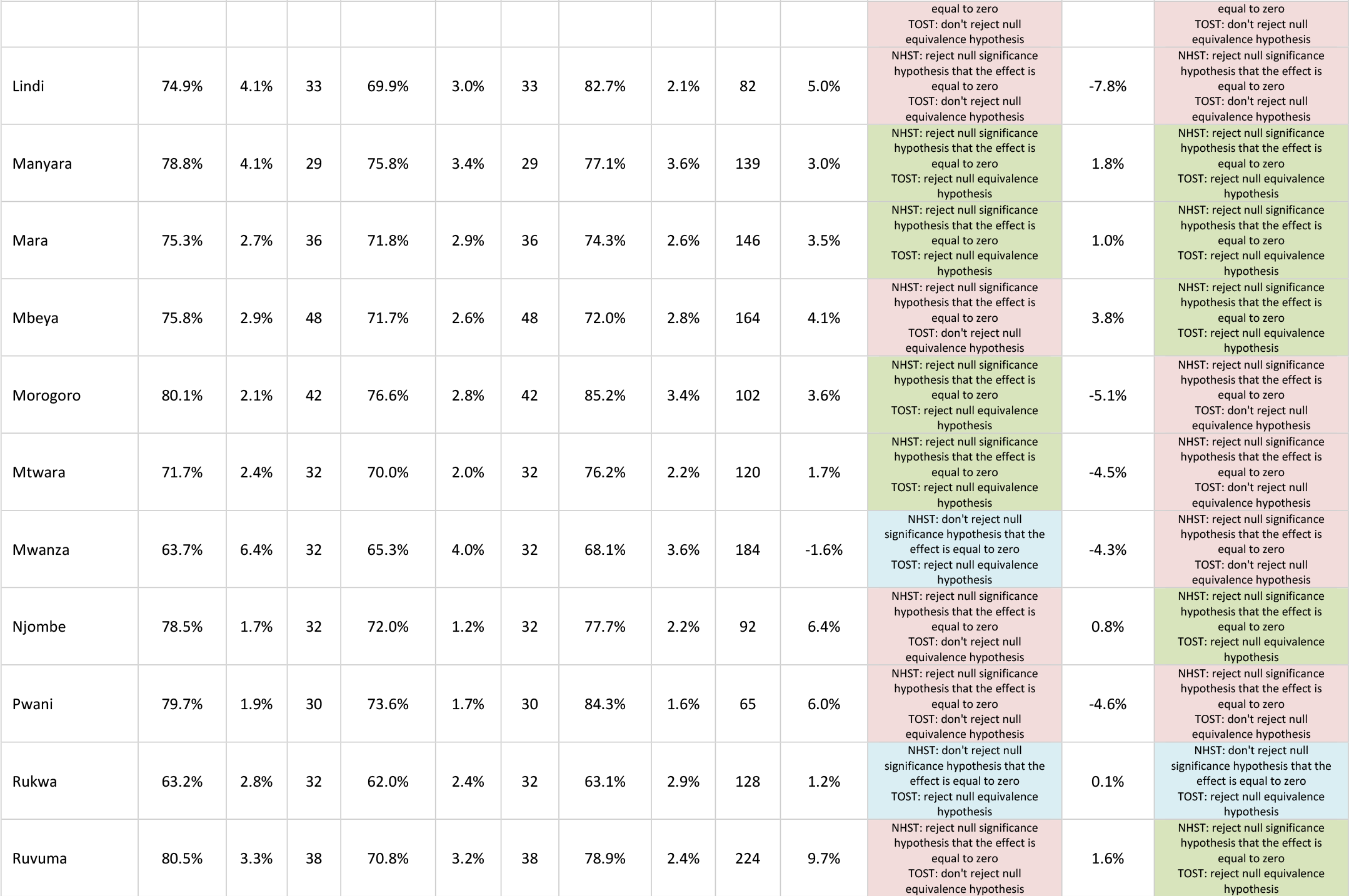
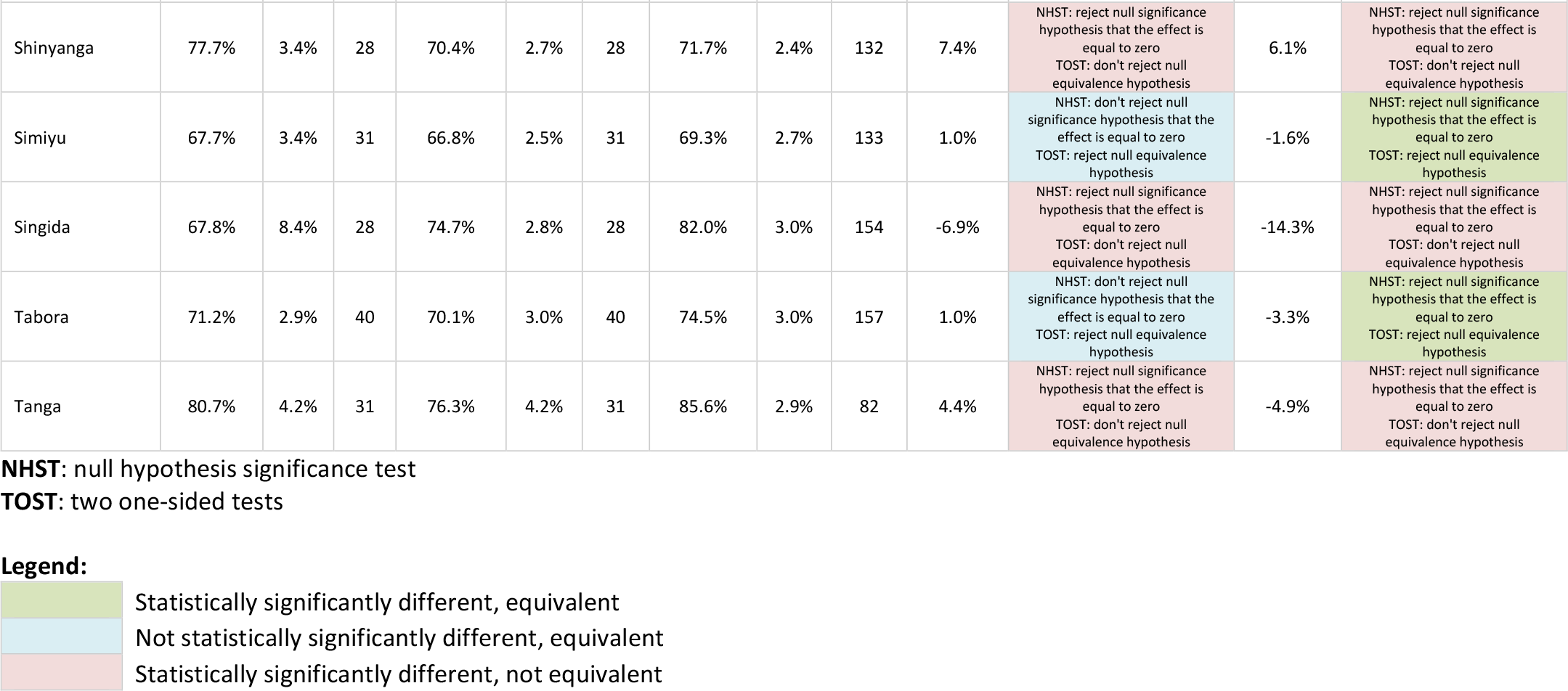
Comparison of caseload-weighted readiness to facility-weighted readiness and observation-weighted readiness, by facility type, managing authority, urban/rural, and region.

|  | Method: caseload-weighted |  |  | Method: facility-weighted |  |  | Method: observation-weighted |  |  | Difference: caseload – facility | Equivalence (bounds -5% and 5%) | Difference: caseload – observation | Equivalence (bounds -5% and 5%) |
| --- | --- | --- | --- | --- | --- | --- | --- | --- | --- | --- | --- | --- | --- |
|  | Estimate | SE | N | Estimate | SE | N | Estimate | SE | N |  |  |  |  |
| National | 75.4% | 1.2% | 860 | 71.8% | 0.6% | 860 | 75.5% | 0.7% | 3466 | 3.7% | NHST: reject null significance hypothesis that the effect is equal to zero<br>TOST: reject null equivalence hypothesis | -0.1% | NHST: reject null significance hypothesis that the effect is equal to zero<br>TOST: reject null equivalence hypothesis |
| **Facility type** |  |  |  |  |  |  |  |  |  |  |  |  |  |
| Hospital | 93.3% | 0.6% | 216 | 90.9% | 1.5% | 216 | 93.3% | 0.5% | 1334 | 2.4% | NHST: reject null significance hypothesis that the effect is equal to zero<br>TOST: reject null equivalence hypothesis | 0.0% | NHST: don't reject null significance hypothesis that the effect is equal to zero<br>TOST: reject null equivalence hypothesis |
| Health center | 84.2% | 0.9% | 310 | 82.3% | 0.6% | 310 | 81.9% | 0.8% | 1274 | 1.8% | NHST: reject null significance hypothesis that the effect is equal to zero<br>TOST: reject null equivalence hypothesis | 2.3% | NHST: reject null significance hypothesis that the effect is equal to zero<br>TOST: reject null equivalence hypothesis |
| Dispensary | 68.8% | 1.5% | 321 | 69.0% | 0.7% | 321 | 68.4% | 1.0% | 840 | -0.2% | NHST: reject null significance hypothesis that the effect is equal to zero<br>TOST: reject null equivalence hypothesis | 0.4% | NHST: reject null significance hypothesis that the effect is equal to zero<br>TOST: reject null equivalence hypothesis |
| Clinic | 88.0% | 2.0% | 13 | 84.7% | 2.5% | 13 | 85.4% | 3.4% | 18 | 3.3% | NHST: reject null significance hypothesis that the effect is equal to zero<br>TOST: reject null equivalence hypothesis | 2.6% | NHST: reject null significance hypothesis that the effect is equal to zero<br>TOST: reject null equivalence hypothesis |
| **Managing authority** |  |  |  |  |  |  |  |  |  |  |  |  |  |
| Government/public | 73.2% | 1.3% | 622 | 69.4% | 0.6% | 622 | 73.2% | 0.8% | 2439 | 3.8% | NHST: reject null significance hypothesis that the effect is equal to zero<br>TOST: reject null equivalence hypothesis | 0.0% | NHST: don't reject null significance hypothesis that the effect is equal to zero<br>TOST: reject null equivalence hypothesis |
| Private-for-profit | 86.6% | 2.9% | 58 | 86.0% | 1.6% | 58 | 86.8% | 2.9% | 202 | 0.7% | NHST: don't reject null significance hypothesis that the effect is equal to zero<br>TOST: reject null equivalence hypothesis | -0.2% | NHST: don't reject null significance hypothesis that the effect is equal to zero<br>TOST: reject null equivalence hypothesis |
| Parastatal | 93.9% | 1.1% | 11 | 89.5% | 4.4% | 11 | 92.6% | 1.7% | 34 | 4.3% | NHST: reject null significance hypothesis that the effect is equal to zero<br>TOST: don't reject null equivalence hypothesis | 1.2% | NHST: reject null significance hypothesis that the effect is equal to zero<br>TOST: reject null equivalence hypothesis |
| Mission/faith-based | 86.1% | 1.8% | 169 | 82.4% | 2.0% | 169 | 84.5% | 2.9% | 791 | 3.7% | NHST: reject null significance hypothesis that the effect is equal to zero<br>TOST: reject null equivalence hypothesis | 1.6% | NHST: reject null significance hypothesis that the effect is equal to zero<br>TOST: reject null equivalence hypothesis |
|  | Estimate | SE | N | Estimate | SE | N | Estimate | SE | N |  |  |  |  |
| Urban/Rural |  |  |  |  |  |  |  |  |  |  |  |  |  |
| Urban | 86.1% | 1.3% | 265 | 82.1% | 1.2% | 265 | 85.8% | 1.4% | 1463 | 4.0% | NHST: reject null significance hypothesis that the effect is equal to zero<br>TOST: reject null equivalence hypothesis | 0.3% | NHST: reject null significance hypothesis that the effect is equal to zero<br>TOST: reject null equivalence hypothesis |
| Rural | 70.8% | 1.4% | 595 | 69.3% | 0.7% | 595 | 71.4% | 0.9% | 2003 | 1.5% | NHST: reject null significance hypothesis that the effect is equal to zero<br>TOST: reject null equivalence hypothesis | -0.6% | NHST: reject null significance hypothesis that the effect is equal to zero<br>TOST: reject null equivalence hypothesis |
| Region |  |  |  |  |  |  |  |  |  |  |  |  |  |
| Arusha | 78.9% | 2.9% | 37 | 72.5% | 2.1% | 37 | 82.5% | 2.4% | 142 | 6.5% | NHST: reject null significance hypothesis that the effect is equal to zero<br>TOST: don't reject null equivalence hypothesis | -3.5% | NHST: reject null significance hypothesis that the effect is equal to zero<br>TOST: reject null equivalence hypothesis |
| Dar es salaam | 86.8% | 2.4% | 43 | 84.0% | 2.3% | 43 | 84.5% | 3.1% | 210 | 2.8% | NHST: reject null significance hypothesis that the effect is equal to zero<br>TOST: reject null equivalence hypothesis | 2.2% | NHST: reject null significance hypothesis that the effect is equal to zero<br>TOST: reject null equivalence hypothesis |
| Dodoma | 77.4% | 3.5% | 39 | 74.5% | 2.5% | 39 | 84.7% | 3.0% | 119 | 2.9% | NHST: reject null significance hypothesis that the effect is equal to zero<br>TOST: reject null equivalence hypothesis | -7.3% | NHST: reject null significance hypothesis that the effect is equal to zero<br>TOST: don't reject null equivalence hypothesis |
| Geita | 77.2% | 2.3% | 27 | 67.6% | 2.2% | 27 | 69.3% | 2.0% | 164 | 9.6% | NHST: reject null significance hypothesis that the effect is equal to zero<br>TOST: don't reject null equivalence hypothesis | 7.9% | NHST: reject null significance hypothesis that the effect is equal to zero<br>TOST: don't reject null equivalence hypothesis |
| Iringa | 76.0% | 3.2% | 30 | 71.8% | 4.3% | 30 | 82.4% | 4.4% | 115 | 4.2% | NHST: reject null significance hypothesis that the effect is equal to zero<br>TOST: don't reject null equivalence hypothesis | -6.5% | NHST: reject null significance hypothesis that the effect is equal to zero<br>TOST: don't reject null equivalence hypothesis |
| Kagera | 74.7% | 2.3% | 40 | 71.5% | 1.9% | 40 | 76.1% | 2.3% | 189 | 3.2% | NHST: reject null significance hypothesis that the effect is equal to zero<br>TOST: reject null equivalence hypothesis | -1.4% | NHST: reject null significance hypothesis that the effect is equal to zero<br>TOST: reject null equivalence hypothesis |
| Katavi | 72.5% | 2.6% | 29 | 68.3% | 2.0% | 29 | 76.2% | 3.0% | 112 | 4.2% | NHST: reject null significance hypothesis that the effect is equal to zero<br>TOST: don't reject null equivalence hypothesis | -3.7% | NHST: reject null significance hypothesis that the effect is equal to zero<br>TOST: reject null equivalence hypothesis |
| Kigoma | 69.5% | 2.2% | 36 | 63.2% | 2.0% | 36 | 65.1% | 2.8% | 197 | 6.4% | NHST: reject null significance hypothesis that the effect is equal to zero<br>TOST: don't reject null equivalence hypothesis | 4.5% | NHST: reject null significance hypothesis that the effect is equal to zero<br>TOST: don't reject null equivalence hypothesis |
| Kilimanjaro | 83.4% | 2.4% | 37 | 74.7% | 3.1% | 37 | 90.8% | 2.2% | 114 | 8.6% | NHST: reject null significance hypothesis that the effect is | -7.5% | NHST: reject null significance hypothesis that the effect is |
|  | Estimate | SE | N | Estimate | SE | N | Estimate | SE | N |  |  |  |  |
|  |  |  |  |  |  |  |  |  |  |  | equal to zero<br>TOST: don't reject null equivalence hypothesis |  | equal to zero<br>TOST: don't reject null equivalence hypothesis |
| Lindi | 74.9% | 4.1% | 33 | 69.9% | 3.0% | 33 | 82.7% | 2.1% | 82 | 5.0% | NHST: reject null significance hypothesis that the effect is equal to zero<br>TOST: don't reject null equivalence hypothesis | -7.8% | NHST: reject null significance hypothesis that the effect is equal to zero<br>TOST: don't reject null equivalence hypothesis |
| Manyara | 78.8% | 4.1% | 29 | 75.8% | 3.4% | 29 | 77.1% | 3.6% | 139 | 3.0% | NHST: reject null significance hypothesis that the effect is equal to zero<br>TOST: reject null equivalence hypothesis | 1.8% | NHST: reject null significance hypothesis that the effect is equal to zero<br>TOST: reject null equivalence hypothesis |
| Mara | 75.3% | 2.7% | 36 | 71.8% | 2.9% | 36 | 74.3% | 2.6% | 146 | 3.5% | NHST: reject null significance hypothesis that the effect is equal to zero<br>TOST: reject null equivalence hypothesis | 1.0% | NHST: reject null significance hypothesis that the effect is equal to zero<br>TOST: reject null equivalence hypothesis |
| Mbeya | 75.8% | 2.9% | 48 | 71.7% | 2.6% | 48 | 72.0% | 2.8% | 164 | 4.1% | NHST: reject null significance hypothesis that the effect is equal to zero<br>TOST: don't reject null equivalence hypothesis | 3.8% | NHST: reject null significance hypothesis that the effect is equal to zero<br>TOST: reject null equivalence hypothesis |
| Morogoro | 80.1% | 2.1% | 42 | 76.6% | 2.8% | 42 | 85.2% | 3.4% | 102 | 3.6% | NHST: reject null significance hypothesis that the effect is equal to zero<br>TOST: reject null equivalence hypothesis | -5.1% | NHST: reject null significance hypothesis that the effect is equal to zero<br>TOST: don't reject null equivalence hypothesis |
| Mtwara | 71.7% | 2.4% | 32 | 70.0% | 2.0% | 32 | 76.2% | 2.2% | 120 | 1.7% | NHST: reject null significance hypothesis that the effect is equal to zero<br>TOST: reject null equivalence hypothesis | -4.5% | NHST: reject null significance hypothesis that the effect is equal to zero<br>TOST: don't reject null equivalence hypothesis |
| Mwanza | 63.7% | 6.4% | 32 | 65.3% | 4.0% | 32 | 68.1% | 3.6% | 184 | -1.6% | NHST: don't reject null significance hypothesis that the effect is equal to zero<br>TOST: reject null equivalence hypothesis | -4.3% | NHST: reject null significance hypothesis that the effect is equal to zero<br>TOST: don't reject null equivalence hypothesis |
| Njombe | 78.5% | 1.7% | 32 | 72.0% | 1.2% | 32 | 77.7% | 2.2% | 92 | 6.4% | NHST: reject null significance hypothesis that the effect is equal to zero<br>TOST: don't reject null equivalence hypothesis | 0.8% | NHST: reject null significance hypothesis that the effect is equal to zero<br>TOST: reject null equivalence hypothesis |
| Pwani | 79.7% | 1.9% | 30 | 73.6% | 1.7% | 30 | 84.3% | 1.6% | 65 | 6.0% | NHST: reject null significance hypothesis that the effect is equal to zero<br>TOST: don't reject null equivalence hypothesis | -4.6% | NHST: reject null significance hypothesis that the effect is equal to zero<br>TOST: don't reject null equivalence hypothesis |
| Rukwa | 63.2% | 2.8% | 32 | 62.0% | 2.4% | 32 | 63.1% | 2.9% | 128 | 1.2% | NHST: don't reject null significance hypothesis that the effect is equal to zero<br>TOST: reject null equivalence hypothesis | 0.1% | NHST: don't reject null significance hypothesis that the effect is equal to zero<br>TOST: reject null equivalence hypothesis |
| Ruvuma | 80.5% | 3.3% | 38 | 70.8% | 3.2% | 38 | 78.9% | 2.4% | 224 | 9.7% | NHST: reject null significance hypothesis that the effect is equal to zero<br>TOST: don't reject null equivalence hypothesis | 1.6% | NHST: reject null significance hypothesis that the effect is equal to zero<br>TOST: reject null equivalence hypothesis |
|  | Estimate | SE | N | Estimate | SE | N | Estimate | SE | N |  |  |  |  |
| Shinyanga | 77.7% | 3.4% | 28 | 70.4% | 2.7% | 28 | 71.7% | 2.4% | 132 | 7.4% | NHST: reject null significance hypothesis that the effect is equal to zero<br>TOST: don't reject null equivalence hypothesis | 6.1% | NHST: reject null significance hypothesis that the effect is equal to zero<br>TOST: don't reject null equivalence hypothesis |
| Simiyu | 67.7% | 3.4% | 31 | 66.8% | 2.5% | 31 | 69.3% | 2.7% | 133 | 1.0% | NHST: don't reject null significance hypothesis that the effect is equal to zero<br>TOST: reject null equivalence hypothesis | -1.6% | NHST: reject null significance hypothesis that the effect is equal to zero<br>TOST: reject null equivalence hypothesis |
| Singida | 67.8% | 8.4% | 28 | 74.7% | 2.8% | 28 | 82.0% | 3.0% | 154 | -6.9% | NHST: reject null significance hypothesis that the effect is equal to zero<br>TOST: don't reject null equivalence hypothesis | -14.3% | NHST: reject null significance hypothesis that the effect is equal to zero<br>TOST: don't reject null equivalence hypothesis |
| Tabora | 71.2% | 2.9% | 40 | 70.1% | 3.0% | 40 | 74.5% | 3.0% | 157 | 1.0% | NHST: don't reject null significance hypothesis that the effect is equal to zero<br>TOST: reject null equivalence hypothesis | -3.3% | NHST: reject null significance hypothesis that the effect is equal to zero<br>TOST: reject null equivalence hypothesis |
| Tanga | 80.7% | 4.2% | 31 | 76.3% | 4.2% | 31 | 85.6% | 2.9% | 82 | 4.4% | NHST: reject null significance hypothesis that the effect is equal to zero<br>TOST: don't reject null equivalence hypothesis | -4.9% | NHST: reject null significance hypothesis that the effect is equal to zero<br>TOST: don't reject null equivalence hypothesis |

NHST: null hypothesis significance test
TOST: two one-sided tests

**Legend:**
|  |  |
| --- | --- |
|  | Statistically significantly different, equivalent |
|  | Not statistically significantly different, equivalent |
|  | Statistically significantly different, not equivalent |

## Notes

### Competing Interest Statement

The authors have declared no competing interest.

## REFERENCES

1. Amouzou A, Leslie HH, Ram M, Fox M, Jiwani SS, Requejo J, et al. Advances in the measurement of coverage for RMNCH and nutrition: from contact to effective coverage. BMJ Glob Health. 2019;4(Suppl 4):e001297. Epub 2019/07/13. doi: 10.1136/bmjgh-2018-001297. PubMed PMID: 31297252; PubMed Central PMCID: PMCPMC6590972.

2. Do M, Micah A, Brondi L, Campbell H, Marchant T, Eisele T, et al. Linking household and facility data for better coverage measures in reproductive, maternal, newborn, and child health care: systematic review. Journal of global health. 2016;6(2):020501. Epub 2016/09/09. doi: 10.7189/jogh.06.020501. PubMed PMID: 27606060; PubMed Central PMCID: PMCPMC5012234.

3. Carter ED, Leslie HH, Marchant T, Amouzou A, Munos MK. Methodological considerations for linking household and healthcare provider data for estimating effective coverage: a systematic review. BMJ open. 2021;11(8). doi: https://doi:10.1136/bmjopen-2020-045704. PubMed PMID: 34446481.

4. Willey B, Waiswa P, Kajjo D, Munos M, Akuze J, Allen E, et al. Linking data sources for measurement of effective coverage in maternal and newborn health: what do we learn from individual-vs ecological-linking methods? Journal of global health. 2018;8(1):010601. Epub 2018/03/03. doi: https://doi:10.7189/jogh.08.010601. PubMed PMID: 29497508; PubMed Central PMCID: PMCPMC5823029 form at https://www.icmje.org/coi_disclosure.pdf (available on request from the corresponding author). Drs. Marchant and Munos report grants from the Bill & Melinda Gates Foundation, and Dr Akuze reports grants from Makerere University School of Public Health during the conduct of the study. All other authors declare no conflict of interest.

5. Munos MK, Maiga A, Do M, Sika GL, Carter ED, Mosso R, et al. Linking household survey and health facility data for effective coverage measures: a comparison of ecological and individual linking methods using the Multiple Indicator Cluster Survey in Côte d’Ivoire. Journal of global health. 2018;8(2):020803. Epub 2018/11/10. doi: 10.7189/jogh.08.020803. PubMed PMID: 30410743; PubMed Central PMCID: PMCPMC6211616 form at https://www.icmje.org/coi_disclosure.pdf (available on request from the corresponding author) and declare no conflict of interest.

6. Tanzania Ministry of Health & Social Welfare, Tanzania National Bureau of Statistics, ICF International. Tanzania Service Provision Assessment Survey 2014-2015. 2016.

7. Tanzania National Bureau of Statistics. Statistical Abstract 2014 2014. Available from: http://www.nbs.go.tz/nbstz/index.php/english/tanzania-abstract/375-statistical-abstract-2014.

8. Sheffel A, Zeger S, Heidkamp R, Munos MK. Development of summary indices of antenatal care service quality in Haiti, Malawi and Tanzania. BMJ Open. 2019;9(12):e032558. doi: https://doi.org/10.1136/bmjopen-2019-032558.

9. Lumley T. survey: analysis of complex survey samples. R package version 4.1.1 ed 2020.

10. R Core Team. R: A Language and Environment for Statistical Computing. Vienna, Austria: R Foundation for Statistical Computing; 2022.

11. Mutale W, Chintu N, Amoroso C, Awoonor-Williams K, Phillips J, Baynes C, et al. Improving health information systems for decision making across five sub-Saharan African countries: Implementation strategies from the African Health Initiative. BMC Health Services Research. 2013;13(Suppl 2):S9–S. doi: 10.1186/1472-6963-13-S2-S9.

12. Lemma S, Janson A, Persson L, Wickremasinghe D, Källestål C. Improving quality and use of routine health information system data in low- and middle-income countries: A scoping review. PLoS One. 2020;15(10):e0239683. Epub 2020/10/09. doi: 10.1371/journal.pone.0239683. PubMed PMID: 33031406; PubMed Central PMCID: PMCPMC7544093.

